# Rapid, Non-Invasive Breath Analysis for Enhancing Detection of Silicosis Using Mass Spectrometry and Interpretable Machine Learning

**DOI:** 10.1101/2024.09.30.24314664

**Authors:** Merryn J Baker, Jeff Gordon, Aruvi Thiruvarudchelvan, Deborah Yates, William A Donald

## Abstract

Occupational lung diseases, such as silicosis, are a significant global health concern, especially with increasing exposure to engineered stone dust. Early detection of silicosis is helpful for preventing disease progression, but existing diagnostic methods, including X-rays, CT scans, and spirometry, often detect the disease only at late stages. This study investigates a rapid, non-invasive diagnostic approach using atmospheric pressure chemical ionization-mass spectrometry (APCI-MS) to analyse volatile organic compounds (VOCs) in exhaled breath from 31 silicosis patients and 60 healthy controls. Six different interpretable machine learning (ML) models with Shapley additive explanations (SHAP) were applied to classify these samples and determine VOC features that contribute the most significantly to model accuracy. The extreme gradient boosting (XGB) classifier demonstrated the best performance, achieving an area under the receiver-operator characteristic curve of 0.933 with the top ten SHAP features. The *m*/*z* 442 feature, potentially corresponding to leukotriene-E3, emerged as a significant predictor for silicosis. The VOC sampling and measurement process takes less than five minutes per sample, highlighting its potential suitability for large-scale population screening. Moreover, the ML models are interpretable through SHAP, providing insights into the features contributing to the model’s predictions. This study suggests that APCI-MS breath analysis could enable early and non-invasive diagnosis of silicosis, helping to improve disease outcomes.

## Introduction

Workplace dust exposures contribute substantially to the burden of occupational lung diseases (OLDs) with a resurgence of pneumoconiosis (or dust-induced lung fibrosis) over the last 10 years.^1^ In particular, silicosis is an emerging epidemic both in Australia and internationally, impacting broad cross-sections of the population.^2–4^ Globally there were over 40,000 deaths attributable to silicosis in 2013, and a recent study found that more than one in four stonemasons in Victoria who had worked with artificial stone benchtops developed silicosis.^5,6^ In 2019, it is estimated that there were 350 cases of silicosis in Australia, largely due to the growth in the engineered stone industry.^7^ Without established treatments, silicosis and silica-related diseases will continue to strain the resources of the health and medical sectors. The development of approaches to identify such diseases early and prevent further exposure can slow the rate of disease progression and allow treatments (once established) to be implemented. While preventative measures exist to reduce exposure (e.g. exposure limits and personal protective equipment), they are often not adhered to; hence early detection has the greatest potential to improve survival rates.^8^

OLDs, including silicosis, are currently detected and diagnosed using long established methods including questionnaires, spirometry and plain chest radiographs.^9^ However, these methods are limited in that they are insensitive for early disease and do not detect silicosis until it has significantly progressed. While chest X-rays are the traditional mainstay of diagnosis and enable classification of disease according to the International Labour Organisation (ILO) system, they are insensitive for detecting disease in early stages as there is a delay between histopathological onset and radiographically visible lesions.^10^ One recent study found that chest X-rays had sensitivities of only 48% when screening for disease presenting with only minimal opacities on the X-ray.^11^ Furthermore, X-ray images cannot definitively identify specific disease due to radiological similarities and clinical overlap with other lung diseases.^12^ Hence, further confirmatory invasive testing such as biopsies may be required for diagnosis. Unlike plain chest radiography, CT scans offer a much higher level of detail and can be used to detect diseases in the early stage, but involve exposure to higher doses of radiation, and are more expensive.^13^ Similarly, spirometry has shown limited sensitivity when detecting abnormalities in lung function in early-stage disease and even in severe disease results may fall within reference ranges.^14^ Current silicosis surveillance methods rely on progression of lung disease to the stage that this is visible in imaging techniques or has significantly restrictive or obstructive lung function below the lower limit of normal. This reduces the likelihood of OLDs being detected in the very early stages where disease progression can more effectively be slowed. This highlights the need for new sensitive detection methods that do not rely on the visibility of the disease and can detect disease in its very early stages. These methods should also ideally be rapid, inexpensive and conducted at the point of care to allow for large scale screening and data gathering.

Analysis of exhaled breath offers an additional non-invasive diagnostic approach.^15–17^ Human breath contains thousands of volatile organic compounds (VOCs), which have been shown to be sensitive biomarkers of many lung diseases.^18–20^ Breath is quick to collect, sensitive and specific for analysis, and highly acceptable to patients. This field has expanded rapidly over the last 20 years, with significant application in lung disease detection, monitoring and screening. To date, exhaled breath analysis has proven application in asthma, chronic obstructive pulmonary disease, lung cancer and occupational lung diseases,^21^ but has not been widely investigated in silicosis.^22^ The presence of diseases such as OLDs change metabolic processes which alter the chemical profiling found in the breath.^18^ These ‘breathprints’ have been found to be specific for different diseases,^23^ suggesting that specific breath tests can be developed. When crystalline silica is deposited in the lungs, macrophages ingesting the dust particles cause an inflammatory response which releases potential biomarkers, altering the abundances and identities of aldehydes, alkanes, and other volatile organic compounds in breath.^24–27^ Similarly, pulmonary fibrosis has been shown to increase the concentrations of a range of chemicals (carbon monoxide, nitric oxide, protein, 3-nitrotyrosine and 8-isoprostane) in exhaled breath condensate^28^ and in exhaled breath.^29^ Such ‘breathprints’ may enable the detection of early disease in individual workers; however, the concentrations of such chemicals are low, which highlights the need for ultra-sensitive methods in chemical analysis.^30^

Here, untargeted atmospheric pressure chemical ionisation-mass spectrometry (APCI-MS) was used to obtain ‘breathprints’ and discover features that could represent breath biomarkers for the early diagnosis of silicosis. APCI-MS is a highly sensitive and rapid analytical technique that allows for the analysis of VOCs in low concentrations. APCI-MS has been commonly used for the analysis of food volatiles during digestion,^31^ but has yet to be widely explored for disease VOCs. Breath from silicosis patients and healthy controls was compared and analysed using multiple machine learning methods to determine diagnostic performance. Key features used by the algorithms in their predictions were also determined using explainable ML using Shapley Additive Explanation,^32,33^ which potentially represent breath biomarkers of silicosis.

## Materials and Methods

### Subjects

A total of 91 patients were recruited for this study and consisted of adult males aged 36 to 79. The study cohort comprised of two groups: 31 patients with clinically diagnosed silicosis, and 60 healthy controls without any documented lung disorders. Patients with a diagnosis of silicosis were recruited prospectively from specialist clinical practice at Holdsworth House Medical Practice (HHMP), Sydney, NSW. The diagnosis of silicosis was made according to acknowledged criteria (Australian National Dust Diseases Taskforce 2022^9,34^). Control patients were recruited prospectively from both HHMP and The University of New South Wales. Control patients had no respiratory conditions and had no previous crystalline silica exposure, and were chosen to ensure age and gender match with silicosis patients. The demographic characteristics, smoking status, medical history and silica exposure history was recorded for all participants. A summary of the participant demographics is shown in **Table 1**. The study was approved by the University of New South Wales Human Research Ethics Committee (HC2203367) in accordance with the National Statement on Ethical Conduct in Human Research (2007) requirements. Written informed consent was received from all participants.

**Table 1:** Summary of patient demographics.

| Characteristic | Silicosis | Healthy control |
| --- | --- | --- |
| Number | 31 | 60 |
| Silicosis category |  |  |
| Simple | 24 | N/A |
| Complicated | 7 |  |
| Average age<sup>a</sup> | 51 ± 12 | 51 ± 11 |
| Gender (% male) | 100 | 100 |
| Smoking status |  |  |
| Non-smokers | 7 | 48 |
| Ex-smokers | 16 | 7 |
| Current smokers | 7 | 5 |
| Vape users | 3<sup>b</sup> | 8<sup>c</sup> |
| Comorbidities<sup>d</sup> |  |  |
| Hypertension | 7 | 7 |
| Heart disease | 5 | 2 |
| Diabetes | 2 | 3 |
| Liver disease | 0 | 0 |
| None of the above | 18 | 53 |
| Other lung diseases<sup>e</sup> |  |  |
| Asthma | 5 | 0 |
| COPD | 8 | 0 |
| Emphysema | 3 | 0 |
| Latent tuberculosis | 2 | 0 |
| Sleep apnoea | 2 | 0 |
| Hypersensitivity pneumonitis | 1 | 0 |
| Chronic pneumothorax | 1 | 0 |
| Chronic bronchitis | 1 | 0 |
| None | 16 | 60 |

<sup>a</sup> ±1 standard deviation
<sup>b</sup> All three vape users are also in the ex-smoker category
<sup>c</sup> Three vape users also currently smoke, and the other five vape users have never smoked
<sup>d</sup> Three people in the silicosis group and five in the control group had multiple comorbidities
<sup>e</sup> Seven people in the silicosis group had multiple other lung diseases
COPD = chronic obstructive pulmonary disease

### Pulmonary function testing

Pulmonary function was assessed *via* spirometry for all (31) silicosis participants as part of their routine medical care around the time of the breath sampling. Lung function parameters including forced vital capacity (FVC), forced expiratory volume in one second (FEV_1_), diffusing capacity for carbon monoxide (DLCO) and total lung capacity (TLC) were measured using an EasyOne Pro (ndd Medical Technologies). All spirometric tests were performed by qualified practitioners following American Thoracic Society (ATS) guidelines. Fractional exhaled nitric oxide (FeNO) readings were obtained for 24 silicosis participants, following ATS guidelines, including the use of nose clips.^35^ The following was the reference range for normal lung function:^36–38^ FCV: greater than 80% predicted, FEV_1_: greater than 80% predicted, FEV_1_/FVC ratio: greater than 0.7, TLC: greater than 80% predicted, DLCO: greater than 60% predicted, FeNO: less than 25 ppb.

### Breath sampling

Breath samples were collected from participants using a 1 L Tedlar bag (Merck Life Science, Australia) fitted with a 0.25” outer diameter tube and threaded valve for sampling. Participants were instructed to rinse their mouths with water prior to sample collection and provide late expiratory breath by discarding the initial fraction of breath and collecting only the final portion of exhaled breath into the bag. Unlike in FeNO sampling, nose clips were not used, consistent with many breath VOC sampling protocols in the literature.^39^ Samples were stored at room temperature in a dark environment until analysis. All samples were analysed on the same day as collection with a maximum time between collection and analysis of six hours.

### Mass spectrometry

Breath samples were analysed by APCI-MS using a custom-made APCI source consisting of a Teflon tube with a high-voltage needle that was equipped to the inlet of a linear trap quadrupole MS (LTQ-XL, Thermo Fisher Scientific). Samples were infused into the APCI-MS at a flow rate of 420 mL/min. Mass spectra were obtained in positive ionization mode by applying potentials of +3.6 kV, 0 V, and +26 V to the corona discharge needle, capillary inlet and tube lens respectively. The capillary inlet temperature was set to 400 ºC. Spectra were acquired using a mass range of 50 to 600 *m/z* for 90 s per sample. For each sample, the spectra were averaged over 60 s, exported as nominal mass, and normalised to the base peak to obtain relative peak abundances as input for machine learning (ML) algorithms to assess diagnostic performance.

### Machine learning

The APCI-MS data was used as input for analysis by six supervised ML algorithms (neural network (NN), extreme gradient boosting (XGB), logistic regression (LR), random forest (RF), linear discriminant analysis (LDA), and support vector machine (SVM)) to classify persons with silicosis from healthy controls. This was implemented using CRANK-MS^32^ in Python (v. 3.8) to determine the diagnostic performance of machine learning algorithms for binary classification. The hyperparameters for each algorithm were optimised using the grid search script in CRANK-MS^32^ (**Table S1**). The results from the ML algorithms were interpreted using SHAP analysis to determine the contribution of each feature to the model prediction.^32,33^

### Performance metrics

For each algorithm, a bootstrap model was used in which the data set was split randomly 100 times into 60% training data and 40% validation data (i.e., 100 “bootstraps”). Each diagnostic performance metric was calculated based on the mean of the 100 bootstrap measurements, and error was calculated as one standard deviation of the mean. For each ML algorithm, accuracy (ACC), precision/positive predictive value (PPV), sensitivity/recall (SN), specificity (SP), *F*1 score, MCC score, and negative predictive value (NPV) were calculated using Equations 1 to 7:

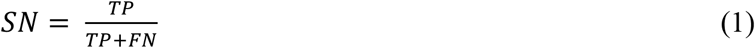

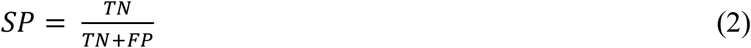

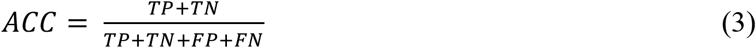

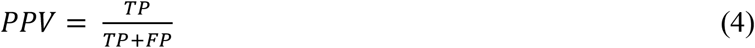

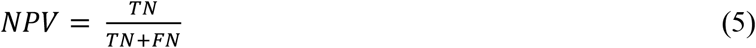

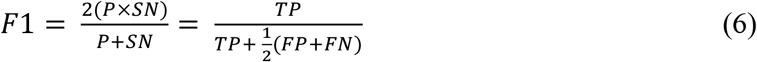

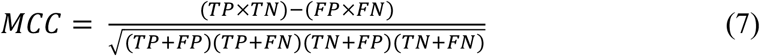

where TP, TN, FP and FN correspond to true positive, true negative, false positive and false negative respectively. Receiver operating characteristic (ROC) and precision-recall (PR) curves were generated and used to calculate area-under-curve (AUC). Briefly, an ROC curve is a plot of the sensitivity against the specificity, and a PR curve is a plot of the precision rate against the sensitivity. These curves are beneficial as they allow for a visual representation of the trade off that often exists between performance metrics, and the area under these curves represent a summary of the model skill. ROC curves are generally more informative when the two classes are balanced in number, and PR curves are preferred when there is an imbalance.

## Results

The characteristics of the silicosis and control groups are outlined in **Table 1**. There were no statistically significant differences between the groups for age and gender. However, the silicosis group exhibited a higher prevalence of comorbidities, including hypertension, heart disease, and diabetes (42% of the silicosis group compared to 12% of the control group), as well as other lung diseases (48% of the silicosis group and 0% of controls, by definition). Additionally, the silicosis group had a higher proportion of both current smokers (23% of the silicosis group compared to 8% of the control group) and ex-smokers (52% of the silicosis group compared to 12% of the control group). Among the silicosis group, 24 participants had simple silicosis, while 7 had the more severe, complicated form. Seventy-seven percent of the silicosis group had been exposed to silica through their occupations as stonemasons or tunnelers, while the remaining participants had been exposed through other professions, such as mining, welding, laboring, and plant operations (**Figure 1d**). Spirometry, diffusion, and FeNO measurements for the silicosis group showed normal readings for most participants (55% to 79%, **Figure 1a-c**), suggesting that most cases were not yet severe enough to cause significant functional impairment.

**Figure 1:**
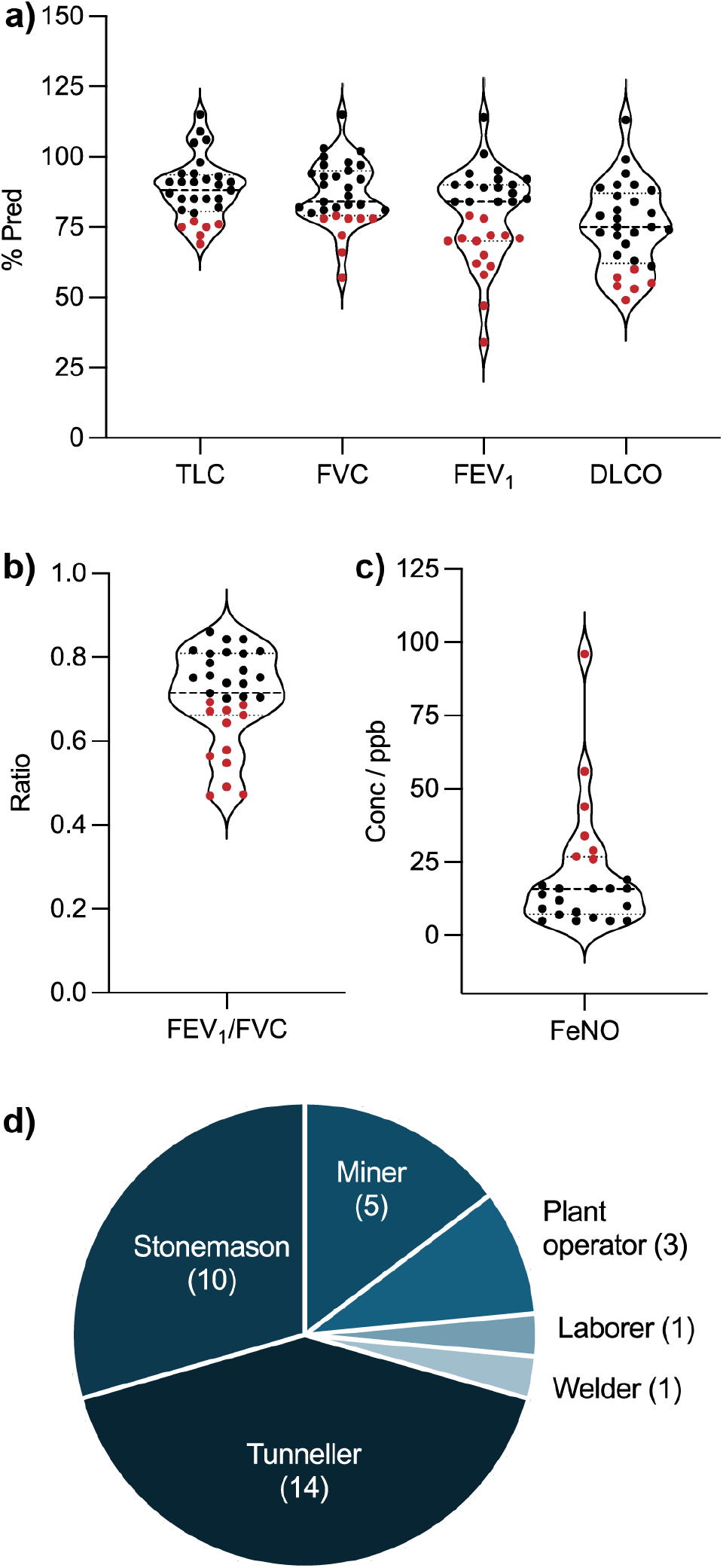
Silicosis group spirometry, diffusion and occupation data. Total lung capacity (TLC), forced vital capacity (FVC), forced expiratory volume in one second (FEV_1_) and diffusing capacity for carbon monoxide (DLCO) are represented as a percentage of predicted (a), and the FEV_1_/FVC ratio and fractional exhaled nitric oxide (FeNO) are represented in panels b) and c). For panels a) to c), data points in black indicate a normal reading and points in red indicate an abnormal reading.^36–38^ Panel d) represents an occupation breakdown for the silicosis patients, noting that three participants worked in multiple industries.

The APCI-MS data from 50 to 600 *m*/*z* was analysed using six machine learning (ML) algorithms to assess their diagnostic performance in classifying silicosis and healthy control samples. Among the evaluated algorithms, the XGB classifier exhibited the highest overall performance, achieving an AUC (ROC) of 0.858 ± 0.055 and an AUC (PR) of 0.760 ± 0.090 (**Figure 2, Table S2**). The LDA and SVM classifiers also showed strong diagnostic capabilities, with AUC (ROC) values of 0.843 ± 0.078 and 0.845 ± 0.061, and AUC (PR) values of 0.756 ± 0.081 and 0.733 ± 0.110, respectively. The performance of the remaining algorithms was similarly robust, with five out of the six classifiers obtaining an AUC (ROC) above 0.8 (**Figure 2, Table S2**).

**Figure 2:**
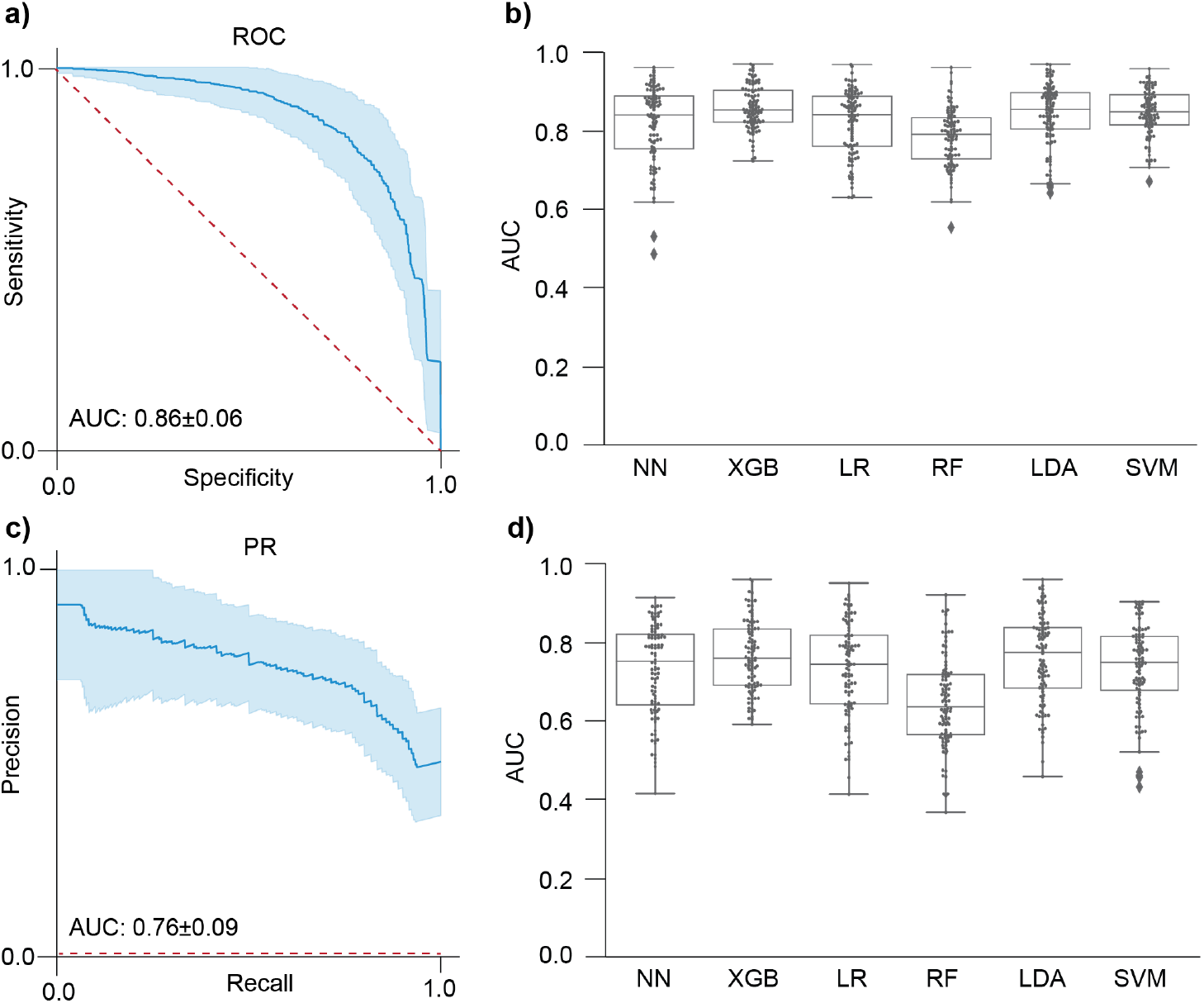
The use of extreme gradient boosting (XGB) outperforms other machine learning algorithms for the classification of Silicosis using APCI-MS data without any feature selection. Receiver-operating characteristic (ROC) curve (a) and precision recall curve (c) for XGB are shown with a 95% CI in blue, and a dotted red line representative of an algorithm with no predictive power. The performance of the six machine learning algorithms is compared through swarm plots of the area under the curve (AUC) for ROC curves (b) and PR curves (d), with points representing each of 100 bootstraps performed for each of the algorithms: neural networks (NN), extreme gradient boost (XGB), logistic regression (LR), random forest (RF), linear discriminant analysis (LDA) and support vector machine (SVM).

The MCC score was highest for the XGB classifier with a normalized score of 0.764 ± 0.178 (**Table S2**). MCC is recognized as a more informative metric for binary classification, particularly in datasets with an imbalance in the size of the cohorts^40^. Other classifiers, including LR, LDA, and SVM, demonstrated slightly lower MCC scores of 0.760 ± 0.189, 0.749 ± 0.224, and 0.742 ± 0.204, respectively (**Table S2**). In terms of the negative predictive value (NPV), the LR classifier performed the best with an NPV of 0.823 ± 0.071, closely followed by the XGB and SVM classifiers with NPVs of 0.814 ± 0.079 and 0.810 ± 0.080, respectively (**Table S2**). NPV is clinically important because it assesses the likelihood that subjects who receive a negative test result are truly healthy, thereby helping to minimize unnecessary follow-up tests. For all six algorithms, randomly permuting the silicosis and healthy data labels resulted in average accuracies that are statistically the same as 50% within one standard deviation (**Figure S1**), consistent with a “random guess” for binary classification as expected for this control test.

To further investigate the contribution of specific features to the classification performance, SHAP analysis was conducted, revealing that the feature at *m*/*z* 442 was the most significant predictor across all six algorithms (**Figure 3**). The *m*/*z* 442 feature intensity was statistically significant between the silicosis and control groups with a *p*-value of 2 x 10^-4^ (**Figure S2, Table S3**). Additional SHAP features, including ions at *m*/*z* 272, 171, and 277, also contributed to the model predictions, though to a lesser extent (**Figure 3**). When the ML algorithms were applied to a reduced dataset containing only the top 26 SHAP features, an improvement in diagnostic performance was observed across all algorithms (**Figure 4a-c, Table S2**), with the XGB classifier achieving an AUC (ROC) of 0.917 ± 0.042 and an AUC (PR) of 0.850 ± 0.080. Further reduction of the feature set to the top ten SHAP features resulted in the highest performance metrics, with the XGB classifier achieving an AUC (ROC) of 0.933 ± 0.038, an AUC (PR) of 0.882 ± 0.073, a normalised MCC score of 0.852 ± 0.126 and a NPV of 0.879 ± 0.069 (**Figure 4d-f, Table S4**). Feature reduction to the top five features and top one feature did not further improve the predictive performance, with AUC (ROC) values of 0.901 ± 0.039 and 0.798 ± 0.060, and AUC (PR) values of 0.859 ± 0.064 and 0.634 ± 0.107, for five and one feature/s, respectively (**Figure 4d-f, Table S4**).

**Figure 3:**
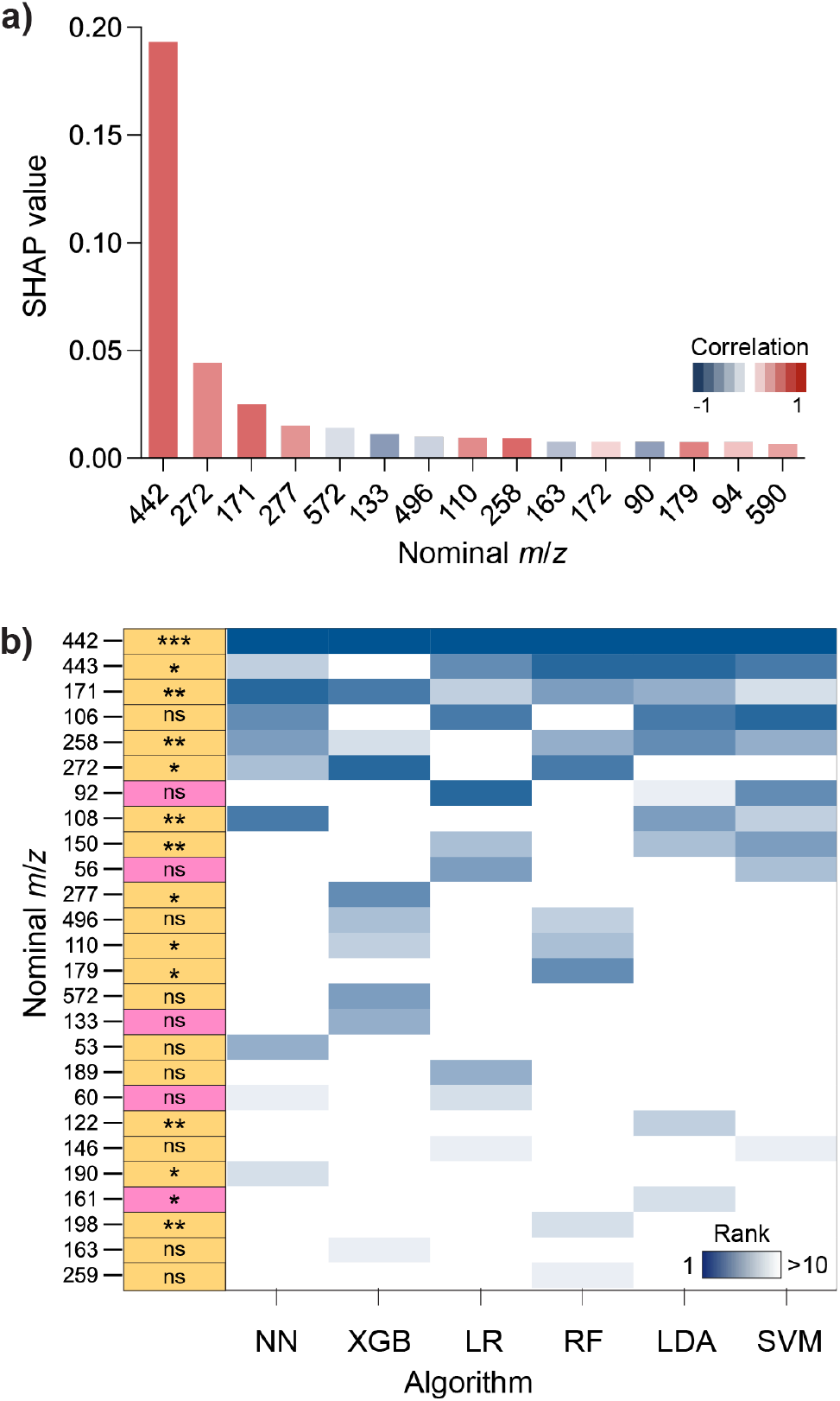
Key nominal *m*/*z* values from APCI-MS analysis of breath used in algorithm predictions. Panel a) shows Shapley additive explanations (SHAP) values for the top 15 ions (nominal *m/*z) that had the highest contribution to a correct silicosis prediction using the XGB classifier. The average correlation corresponds to whether the feature is greater in intensity (red) or lower in intensity (blue) in silicosis samples. Panel b) shows comparative SHAP rankings for the top 10 metabolites for all six algorithms with the first column summarising statistical analysis of the features including P-value (P < 0.0001 = ***, P < 0.001 = **, P < 0.05 = * and P > 0.05 = not significant (ns)) and fold change relative to healthy (yellow = higher intensity in silicosis, pink = higher intensity in healthy).

**Figure 4:**
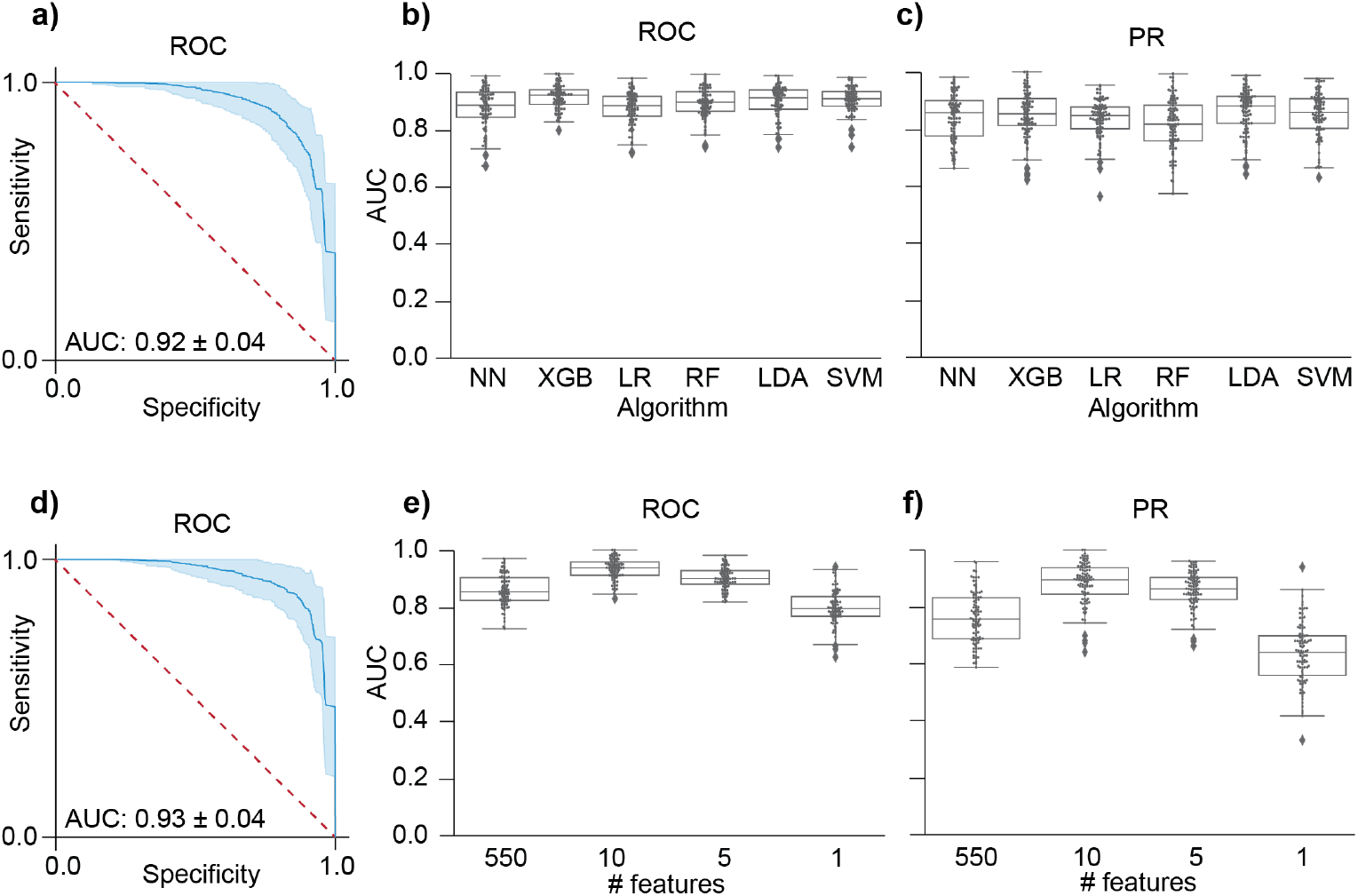
Using a feature-selected model based on SHAP rank increased the performance of all algorithms with XGB remaining the best performing classifier. Panels a) to c) represent algorithm performance using the combined top 10 SHAP features from all algorithms (26 features total), with a) showing the ROC curve for XGB classifier, and b) and c) showing comparative swarm plots of the AUC for ROC curves and PR curves, respectively. Panels d) to f) represent a comparison of the performance of XGB with differing number of features. The ROC curve for the XGB classifier using the top 10 SHAP features is shown in a), and comparative swarm plots of AUC for ROC and PR curves for differing numbers of input features are shown in e) and f), respectively.

## Discussion

This study aimed to evaluate the diagnostic performance of various machine learning (ML) algorithms in classifying silicosis and healthy control samples using non-invasive breath sampling coupled with a rapid VOC fingerprinting test through APCI-MS, rather than targeting specific VOCs. The results demonstrated that the XGB classifier consistently outperformed other algorithms across most performance metrics, particularly in terms of ROC and PR AUC values, and normalised MCC score (0.933 ± 0.038, 0.882 ± 0.073 and 0.852 ± 0.126, respectively using the top ten SHAP features). These findings suggest that the overall approach, particularly when using the XGB classifier, can be highly effective in facilitating the rapid and accurate diagnosis of silicosis, owing to its ability to account for complex patterns in the VOC data that may not be readily apparent with alternative methods.

Breath tests have emerged as a promising non-invasive tool for diagnosing various respiratory conditions, including chronic obstructive pulmonary disease (COPD)^41^ and pneumoconiosis.^23^ Most of the current research in this area has relied on gas chromatography-mass spectrometry (GC-MS) to identify volatile organic compounds (VOCs) in exhaled breath that can serve as biomarkers for these diseases. While GC-MS is highly sensitive and capable of detailed chemical analysis, it typically involves long analysis times, making it less practical for large-scale screening efforts. For COPD, studies using GC-MS have shown that specific VOC profiles can differentiate between disease stages^42^ and even predict exacerbations^43^. Similarly, breath analysis in asbestosis has identified compounds linked to lung inflammation and fibrosis,^44^ though the specificity of these biomarkers remains a challenge. Despite these advances, only two studies to date have examined the potential of breath analysis for silicosis diagnosis.^22,45^ Both studies used solid phase microextraction (SPME) GC-MS for analysis. One study from 2016 only included four silicosis participants, and hence could not make conclusions on the diagnostic performance of the identified potential biomarkers.^45^ A study from 2023 with greater patient numbers also identified potential biomarkers using orthogonal partial least squares discriminant analysis, obtaining a sensitivity and specificity of 60.3% and 89.2%.^22^ While the use of SPME-GC-MS is beneficial in allowing for more detailed compound identification, the total extraction and run time for each sample is 95 minutes, which presents a challenge for using it widely as a screening tool. For example, Coal Services Australia conducted nearly 9792 screening tests in the 2022-23 reporting year,^46^ which would require over 15,500 hours of sampling processing and instrument time if exhaled breath analysis was included in the screening process.

The approach used in this study, utilizing APCI-MS, offers a significant advantage with an analysis time of less than two minutes and no preconcentration is required. This rapid analysis opens the possibility of its application in larger-scale screening including routine respiratory surveillance, which is particularly relevant for silicosis given its irreversible nature and lack of effective treatments. Early and accurate diagnosis through such a test could be crucial in early diagnosis and prevention of disease progression by removal from further exposure, also potentially reducing the reliance on invasive procedures. However, further confirmation of specificity and sensitivity is required in a prospective double-blind study using larger numbers of participants.

This study represents a potential advance in respiratory disease diagnostics, as it the first example in the literature of a rapid, non-invasive breath test for silicosis that is feasible for large-scale screening. Traditionally, silicosis has been diagnosed through a combination of clinical assessment, imaging studies such as chest X-rays or CT scans, and pulmonary function tests, all of which present challenges in terms of accuracy and specificity. For instance, radiography modalities have sensitivity ranges between 48% and 68%,^11,47^ with potential overlap in findings with other lung conditions, while pulmonary function tests may only detect changes in advanced stages of the disease, with reported positive predictive values of 41% to 58%.^48^ This trend is similarly observed in this study, where 55% to 79% of spirometry and diffusion measures were in the normal range for silicosis participants. In some cases, invasive procedures like bronchoscopy or lung biopsy are required. By leveraging advanced machine learning algorithms to analyse breath VOC profiles obtained through APCI-MS and using them in addition to conventional testing, this research suggests that exhaled breath profiling could represent promising rapid and non-invasive biomarker analysis to facilitate the diagnosis of silicosis.

The APCI process used for sample analysis results in the formation of protonated monomers [M+H]^+^, with a nominal neutral mass of 441 Da corresponding to the 442 *m*/*z* feature, tentatively assigned to leukotriene-E3 (LTE3), a molecule with an exact mass of 441.25 Da. While leukotrienes, including LTE4, LTD4, and LTB4, are well-established markers in obstructive lung diseases such as asthma^49^ and chronic obstructive pulmonary disease (COPD)^50^ their presence in restrictive lung diseases like silicosis has been less explored. Recent evidence suggests that leukotrienes could play a role in fibrotic lung diseases through their involvement in inflammatory signalling pathways. Elevated levels of LTE4 and LTD4 have been observed in pneumoconiosis linked to asbestos and silica exposure.^51^ Given the inflammatory nature of silicosis, driven largely by the NLRP3 inflammasome, it is plausible that leukotrienes, which are potent mediators of inflammation, could also be involved in the disease process.^52^ Leukotriene antagonists, commonly used in asthma, are not typically applied in silicosis, as this disease involves different inflammatory pathways, particularly those regulated by the NLRP3 inflammasome rather than leukotriene-mediated mechanisms.^53,54^ Thus, its potential as a biomarker for silicosis and an understanding of its interaction with NLRP3-mediated inflammation should be further explored.

### Limitations

While the results are promising, several limitations are acknowledged. First, the study’s sample size is relatively small, and expanding the population to include a broader range of lung conditions and stages of silicosis is necessary. This is particularly important for differentiating silicosis from other similar respiratory conditions such as interstitial pulmonary fibrosis^55^ and sarcoidosis,^56^ which often present overlapping symptoms. Differentiating these diseases is crucial because they may require distinct treatment approaches, and misdiagnosis can lead to inappropriate therapies, particularly for future therapies that are currently in the development pipeline.^55^ Second, unambiguous identification of the detected biomarkers requires further validation. Higher-resolution mass spectrometry and ion fragmentation data, supported by authentic standards, are essential to accurately pinpoint the biomarkers specific to silicosis. This step is critical to confirm the findings and enhance the reliability of these potential diagnostic markers. Future studies should aim to address these gaps to refine the diagnostic utility of the identified biomarkers for silicosis.

## Conclusions

This study provides early evidence for the potential of exhaled breath analysis using APCI-MS as a rapid, non-invasive diagnostic tool for silicosis. The VOC measurement takes less than two minutes per sample and does not require a preconcentration sample processing step, making it a promising candidate for large population screening, offering a significant advantage over methods that are invasive and time-consuming particularly in at-risk occupational groups. The interpretability of the machine learning models, made possible through SHAP provides a transparent way to understand how each feature contributes to the model’s predictions, ensuring that the diagnostic results can be interpreted. For example, the analysis identified the *m*/*z* 442 feature as a significant predictor across multiple machine learning models, which potentially corresponds to leukotriene-E3, a molecule from a class that has been previously linked to lung inflammation. This finding suggests that the *m*/*z* 442 feature could serve as a biomarker for silicosis, although further validation is required to confirm its role. This research highlights the potential of breath-based diagnostics, particularly when combined with interpretable machine learning, to advance early silicosis detection and potentially improve patient outcomes.

## Data Availability

All data produced in the present study are available upon reasonable request to the authors

## Acknowledgements

This work was funded by the iCare Dust Diseases Board through a Discovery and Innovation grant. We thank the specialists and doctors at Holdsworth House Medical Practice for their assistance in recruiting both silicosis and control participants. We thank Nicholas Olsen (Stats Central, UNSW Sydney) for helpful discussions.

## Ethical Statement

The study was approved by the University of New South Wales Human Research Ethics Committee (HC2203367) in accordance with the National Statement on Ethical Conduct in Human Research (2007) requirements. Written informed consent was received from all participants.

