## Supporting Information for "Rapid, Non-Invasive Breath Analysis for Enhancing Detection of Silicosis Using Mass Spectrometry and Interpretable Machine Learning"

**Table of contents**

**Figure S1.** AUC ROC from random label permutations……………………S3

**Table S1**. Algorithm hyperparameters…………………………………...S4

**Table S2.** Diagnostic performance metrics all algorithms…………………….S5

**Figure S2.** Violin plot of 442 *m*/*z*……………………………………...S6

**Table S3.** Statistical summary of key SHAP features………………………... S7

**Table S4.** Diagnostic performance metrics for XGB reduced features.............S8

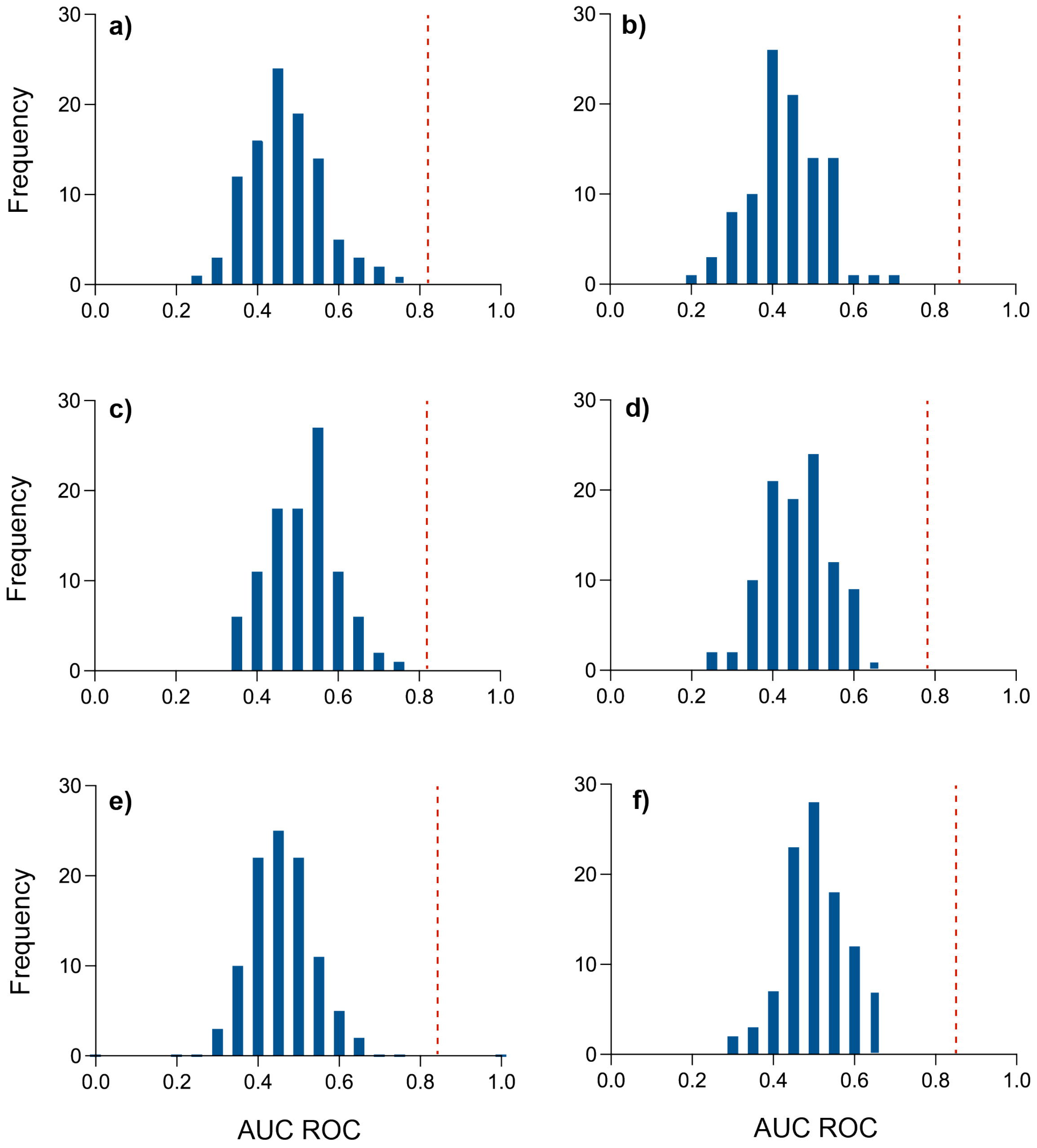


**Figure S1:** Randomly permutating the silicosis and healthy disease labels results in average AUC ROC values (N=100) that are statistically the same as 0.5 within 1 standard deviation. An AUC ROC value of 0.5 corresponds to that obtained by 'random guessing' which is equivalent to a model for binary classification with no predictive power. Histograms of accuracy scores (N = 100) obtained for APCI-MS dataset using NN (a), XGB (b), LR (c), RF (d), LDA (e), and SVM (f) classifiers. The red dashed lines indicate the average accuracy for the original dataset with correct disease label.

**Table S1:** Hyperparameter values applied to each algorithm determined by GridSearch optimisation tuning on the dataset containing all features.

| Algorithm | Hyperparameter |
|---|---|
| Extreme gradient boosting | booster = 'gbtree'<br>learning_rate = 0.5<br>subsample = 1<br>colsample_bytree = 0.6<br>max_depth = 1<br>reg_alpha = 0.6<br>reg_lambda = 0.4<br>eval_metric = 'logloss'<br>n_estimators = 100 |
| Linear discriminant analysis | solver = 'eigen'<br>shrinkage = 'auto' |
| Logistic regression | C = 121,<br>penalty = 'l2'<br>solver = 'liblinear'<br>random_state = seed |
| Neural network | dense_dims = 32<br>num_epochs = 64<br>batch_size = 16<br>lambda_l1 = 0.0005<br>lambda_l2 = 0.007 |
| Random forest | criterion='gini',<br>n_estimators = 25<br>max_depth = None<br>max_features = 'auto'<br>min_samples_split = 2<br>random_state = seed |
| Support vector machine | kernel = 'rbf'<br>C = 50<br>gamma = 0.0005<br>random_state = seed |

**Table S2:** Summary of diagnostic performance metrics for each algorithm and analysed dataset.

| Dataset | NN | XGB | LR | RF | LDA | SVM |
|---|---|---|---|---|---|---|
| All features (550) | Accuracy: 0.7597 ± 0.0690<br>Precision: 0.6943 ± 0.1433<br>Sensitivity: 0.5625 ± 0.1633<br>Specificity: 0.8678 ± 0.0741<br>F1 score: 0.6043 ± 0.1196<br>MCC: 0.4556 ± 0.1557<br>AUC (ROC): 0.8177 ± 0.0974<br>AUC (PR): 0.7274 ± 0.1175<br>NPV: 0.7931 ± 0.0843 | Accuracy: 0.7884 ± 0.0592<br>Precision: 0.7490 ± 0.1278<br>Sensitivity: 0.6126 ± 0.1513<br>Specificity: 0.8882 ± 0.0679<br>F1 score: 0.6547 ± 0.1022<br>MCC: 0.5279 ± 0.1230<br>AUC (ROC): 0.8578 ± 0.0547<br>AUC (PR): 0.7603 ± 0.0902<br>NPV: 0.8144 ± 0.0789 | Accuracy: 0.7870 ± 0.0595<br>Precision: 0.7168 ± 0.1201<br>Sensitivity: 0.6405 ± 0.1344<br>Specificity: 0.8656 ± 0.0675<br>F1 score: 0.6649 ± 0.0998<br>MCC: 0.5209 ± 0.1296<br>AUC (ROC): 0.8209 ± 0.0829<br>AUC (PR): 0.7300 ± 0.1196<br>NPV: 0.8227 ± 0.0711 | Accuracy: 0.7138 ± 0.0659<br>Precision: 0.6554 ± 0.1738<br>Sensitivity: 0.4241 ± 0.1540<br>Specificity: 0.8744 ± 0.0834<br>F1 score: 0.4883 ± 0.1214<br>MCC: 0.3406 ± 0.1438<br>AUC (ROC): 0.7824 ± 0.0737<br>AUC (PR): 0.6434 ± 0.1156<br>NPV: 0.7450 ± 0.0819 | Accuracy: 0.7811 ± 0.0667<br>Precision: 0.7591 ± 0.1341<br>Sensitivity: 0.5456 ± 0.1657<br>Specificity: 0.9079 ± 0.0588<br>F1 score: 0.6169 ± 0.1309<br>MCC: 0.4976 ± 0.1486<br>AUC (ROC): 0.8432 ± 0.0779<br>AUC (PR): 0.7556 ± 0.1125<br>NPV: 0.7945 ± 0.0805 | Accuracy: 0.7705 ± 0.0588<br>Precision: 0.6995 ± 0.1235<br>Sensitivity: 0.6049 ± 0.1649<br>Specificity: 0.8609 ± 0.0700<br>F1 score: 0.6309 ± 0.1110<br>MCC: 0.4844 ± 0.1330<br>AUC (ROC): 0.8445 ± 0.0605<br>AUC (PR): 0.7329 ± 0.1101<br>NPV: 0.8097 ± 0.0798 |
| Top SHAP features (26) | Accuracy: 0.8176 ± 0.0598<br>Precision: 0.7838 ± 0.1190<br>Sensitivity: 0.6744 ± 0.1549<br>Specificity: 0.8987 ± 0.0656<br>F1 score: 0.7084 ± 0.0972<br>MCC: 0.5962 ± 0.1257<br>AUC (ROC): 0.8804 ± 0.0715<br>AUC (PR): 0.8393 ± 0.0801<br>NPV: 0.8414 ± 0.0811 | Accuracy: 0.8389 ± 0.0567<br>Precision: 0.8140 ± 0.1105<br>Sensitivity: 0.7057 ± 0.1270<br>Specificity: 0.9129 ± 0.0578<br>F1 score: 0.7442 ± 0.0897<br>MCC: 0.6418 ± 0.1144<br>AUC (ROC): 0.9171 ± 0.0415<br>AUC (PR): 0.8496 ± 0.0795<br>NPV: 0.8555 ± 0.0706 | Accuracy: 0.8100 ± 0.0557<br>Precision: 0.7530 ± 0.1095<br>Sensitivity: 0.6859 ± 0.1397<br>Specificity: 0.8784 ± 0.0666<br>F1 score: 0.7052 ± 0.0853<br>MCC: 0.5782 ± 0.1196<br>AUC (ROC): 0.8800 ± 0.0557<br>AUC (PR): 0.8338 ± 0.0673<br>NPV: 0.8430 ± 0.0741 | Accuracy: 0.8127 ± 0.0631<br>Precision: 0.7963 ± 0.1266<br>Sensitivity: 0.6351 ± 0.1573<br>Specificity: 0.9122 ± 0.0593<br>F1 score: 0.6888 ± 0.1115<br>MCC: 0.5813 ± 0.1322<br>AUC (ROC): 0.8991 ± 0.0496<br>AUC (PR): 0.8133 ± 0.0899<br>NPV: 0.8277 ± 0.0807 | Accuracy: 0.8300 ± 0.0580<br>Precision: 0.8544 ± 0.1106<br>Sensitivity: 0.6197 ± 0.1358<br>Specificity: 0.9419 ± 0.0481<br>F1 score: 0.7059 ± 0.1018<br>MCC: 0.6162 ± 0.1180<br>AUC (ROC): 0.9027 ± 0.0549<br>AUC (PR): 0.8605 ± 0.0770<br>NPV: 0.8270 ± 0.0706 | Accuracy: 0.8178 ± 0.0647<br>Precision: 0.8488 ± 0.1077<br>Sensitivity: 0.5952 ± 0.1547<br>Specificity: 0.9391 ± 0.0510<br>F1 score: 0.6807 ± 0.1178<br>MCC: 0.5922 ± 0.1187<br>AUC (ROC): 0.9106 ± 0.0422<br>AUC (PR): 0.8481 ± 0.0780<br>NPV: 0.8168 ± 0.0791 |

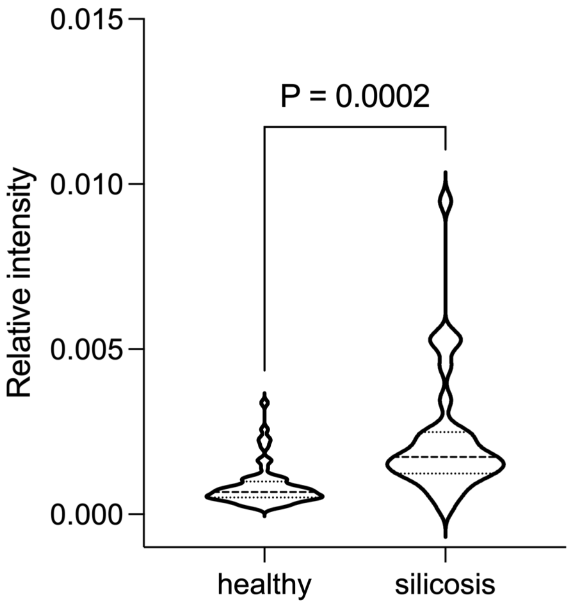


**Figure S2:** Violin plot of the relative intensities of the nominal 442 *m*/*z* feature in silicosis and healthy control samples. P-value calculated from a two-tailed heteroscedastic t-test.

**Table S3:** Statistical summary of collective top ten SHAP features from each algorithm (26 features total, ordered by decreasing average SHAP rank). P-values were calculated using a two-tailed heteroscedastic t-test. Fold change represents the average intensity in silicosis samples divided by the average intensity in healthy samples.

| Nominal m/z | P-value | Fold change |
|---|---|---|
| 442 | $2.0 \times 10^{-4}$ | 2.7 |
| 443 | $3.6 \times 10^{-2}$ | 2.7 |
| 171 | $2.2 \times 10^{-3}$ | 2.5 |
| 106 | $2.6 \times 10^{-1}$ | 1.1 |
| 258 | $6.3 \times 10^{-3}$ | 3.3 |
| 272 | $2.8 \times 10^{-2}$ | 1.9 |
| 92 | $3.6 \times 10^{-1}$ | 0.9 |
| 108 | $2.4 \times 10^{-3}$ | 2.2 |
| 150 | $8.9 \times 10^{-3}$ | 1.6 |
| 56 | $1.6 \times 10^{-1}$ | 0.8 |
| 277 | $1.9 \times 10^{-2}$ | 2.4 |
| 496 | $4.4 \times 10^{-1}$ | 2.8 |
| 110 | $1.5 \times 10^{-2}$ | 3.4 |
| 179 | $1.4 \times 10^{-2}$ | 1.8 |
| 572 | $3.2 \times 10^{-1}$ | 8.7 |
| 133 | $2.3 \times 10^{-1}$ | 0.8 |
| 53 | $9.9 \times 10^{-1}$ | 1.0 |
| 189 | $3.6 \times 10^{-1}$ | 1.1 |
| 60 | $4.8 \times 10^{-1}$ | 0.8 |
| 122 | $3.8 \times 10^{-3}$ | 2.1 |
| 146 | $1.7 \times 10^{-1}$ | 1.4 |
| 190 | $3.2 \times 10^{-2}$ | 1.4 |
| 161 | $1.6 \times 10^{-2}$ | 0.7 |
| 198 | $5.0 \times 10^{-3}$ | 2.0 |
| 163 | $3.5 \times 10^{-1}$ | 1.6 |
| 259 | $5.4 \times 10^{-2}$ | 2.8 |

**Table S4:** Summary of diagnostic performance metrics using the XGB algorithm with differing numbers of input features.

| Dataset | XGB performance metrics | |
|---|---|---|
| Top 10 SHAP features | Accuracy: | 0.8662 ± 0.0512 |
| | Precision: | 0.8488 ± 0.0979 |
| | Sensitivity: | 0.7571 ± 0.1255 |
| | Specificity: | 0.9277 ± 0.0493 |
| | F1 score: | 0.7898 ± 0.0812 |
| | MCC: | 0.7043 ± 0.1038 |
| | AUC (ROC): | 0.9328 ± 0.0375 |
| | AUC (PR): | 0.8821 ± 0.0732 |
| | NPV: | 0.8789 ± 0.0687 |
| Top 5 SHAP features | Accuracy: | 0.8249 ± 0.0498 |
| | Precision: | 0.7788 ± 0.1255 |
| | Sensitivity: | 0.6990 ± 0.1387 |
| | Specificity: | 0.8948 ± 0.0632 |
| | F1 score: | 0.7222 ± 0.0964 |
| | MCC: | 0.6105 ± 0.1145 |
| | AUC (ROC): | 0.9008 ± 0.0386 |
| | AUC (PR): | 0.8594 ± 0.0643 |
| | NPV: | 0.8532 ± 0.0674 |
| Top 1 SHAP feature | Accuracy: | 0.7930 ± 0.0507 |
| | Precision: | 0.6990 ± 0.1062 |
| | Sensitivity: | 0.7130 ± 0.1352 |
| | Specificity: | 0.8391 ± 0.0719 |
| | F1 score: | 0.6937 ± 0.0855 |
| | MCC: | 0.5494 ± 0.1101 |
| | AUC (ROC): | 0.7975 ± 0.0598 |
| | AUC (PR): | 0.6344 ± 0.1071 |
| | NPV: | 0.8502 ± 0.0699 |
